# Differential expression of Angiotensin-Converting Enzyme 2 in Nasal Tissue of Patients with Chronic Rhinosinusitis with Nasal Polyps

**DOI:** 10.1101/2021.02.01.21250623

**Authors:** Philippa C. Fowler, Åsa Torinsson Naluai, Martin Oscarsson, Sara Torkzadeh, Anton Bohman, Mats Bende, Ali M. Harandi

**Affiliations:** Department of Laboratory Medicine, Institute of Biomedicine, Sahlgrenska Academy at the University of Gothenburg, Sweden; Department of Otorhinolaryngology, Skaraborg Hospital, Skövde, Sweden; Department of Otorhinolaryngology, Uppsala University Hospital, Uppsala, Sweden; Department of Microbiology and Immunology, Institute of Biomedicine, Sahlgrenska Academy at the University of Gothenburg, Gothenburg, Sweden; Vaccine Evaluation Center, BC Children’s Hospital Research Institute & Department of Pediatrics, The University of British Columbia, Vancouver, Canada

## Abstract

The coronavirus disease 2019 (COVID-19) caused by the severe acute respiratory syndrome coronavirus 2 (SARS-CoV-2) became a pandemic and a global health emergency. The SARS-CoV-2 receptor angiotensin-converting enzyme 2 (ACE2) is highly expressed in nasal epithelial cells and plays a major role in cellular entry leading to infection. High expression of ACE2 has been suggested to be a potential risk factor for virus infection and disease severity. However the profile of *ACE2* gene expression in diseases of the upper airways remains poorly understood. We herein investigated *ACE2* gene expression in the nasal tissues of a cohort of Swedish patients with chronic rhinosinusitis with nasal polyps (CRSwNPs) using RT-qPCR. *ACE2* mRNA expression was significantly reduced in the nasal mucosa of CRSwNP patients compared to that of controls. Moreover, we observed a sex-dependant difference in nasal *ACE2* expression, where significantly lower levels of the *ACE2* transcript were detected in the nasal mucosa of only female CRSwNP patients. These findings indicate that CRSwNP patients with a decrease in *ACE2* gene expression may thereby be less prone to be infected by SARS-CoV-2. These results enhance our understanding on the profile of *ACE2* expression in the nasal mucosa of patients with upper airway diseases, and their susceptibility to infection with SARS-CoV-2.

---

The coronavirus disease 2019 (COVID-19) is caused by the severe acute respiratory syndrome coronavirus 2 (SARS-CoV-2). Since the detection of the virus in Wuhan, China, COVID-19 has spread worldwide, and the high transmissibility of the virus along with the high fatality rate of the disease has caused a global health emergency [1,2]. Whilst the most common symptoms of patients infected with SARS-CoV-2 include fever and cough, a proportion of patients additionally exhibit multi-organ damage and dysfunction including acute respiratory distress syndrome, acute respiratory injury and acute renal injury [3]. Another very specific symptom which has been established for COVID-19 includes the loss of smell [4]. Further, patients with comorbidities such as hypertension, pulmonary disease and diabetes are highly represented among hospitalized patients with COVID-19 [5,6]. This suggests that the presence of risk factors may determine the susceptibility to SARS-CoV-2 infection. There is hence an urgent need to learn more about the mechanisms involved in human infection and the factors that determine disease severity in patients.

SARS-CoV-2 is a positive sense, single stranded ribonucleic acid enveloped virus comprising of glycoprotein spike (S) on its outer surface which mediates its entry into the cell [7]. It has recently been shown that SARS-CoV-2 uses the angiotensin-converting enzyme 2 (ACE2) as its receptor to gain entry into the cell [7]. In order to fully understand the susceptibility for SARS-CoV-2 infection and the role of ACE2 in the disease severity, a major focus has therefore been aimed at uncovering the cell type-specific gene expression of ACE2 in human tissues. Several studies have recently reported on the gene expression of ACE2 in the respiratory tract. A study of infected COVID-19 patients documented a significantly higher SARS-CoV-2 viral load in nasal swab samples compared to those of throat swabs [8]. Higher ACE2 gene expression has been reported in the nasal compared to throat tissue [9], where ACE2 has been shown to be more highly expressed in the upper respiratory tract compared to the lower, implicating the upper respiratory epithelium as the most likely port of entry for SARS-CoV-2 [10]. Moreover, whereas ACE2 has been shown to be expressed in the epithelial support and stem cells [11], it has also been shown to be co-expressed in the nasal epithelium with genes involved in innate immunity, referring to the potential role of these cells in initiating SARS-CoV-2 infection [12]. Whilst higher ACE2 expression in the upper respiratory epithelium suggests that these cells can serve as a reservoir for viral invasion and facilitate SARS-CoV-2 replication, it is not clear whether any potential underlying factors may affect the expression of ACE2 receptors and therefore increase the severity of the disease. An increased expression of ACE2 has recently been reported in the lungs of patients with comorbidities, suggesting that patients with comorbidities may have a higher risk of developing severe COVID-19 by facilitating SARS-Cov-2 entry into the lungs during infection [13]. Moreover, the infection mediated by SARS-CoV-2 has been shown to increase in severity with age [14,15], where an age-dependant expression of ACE2 has been reported in the nasal epithelium, with lower ACE2 expression found in children which increases with age [16]. Herein, we sought to investigate whether chronic rhinosinusitis with nasal polyps (CRSwNP), influences the expression of ACE2 in the nasal tissues and hence might influence the risk of infectivity with SARS-CoV-2.

CRSwNP is defined as a subgroup of chronic rhinosinusitis (CRS) that is characterised by the presence of fleshy swellings (nasal polyps) of the nasal mucosa, developing in the lining of the nose and paranasal sinuses, presumably due to chronic inflammation. CRSwNP is difficult to treat and recurrences are frequent, despite medical treatment and surgical interventions. Additionally, patients suffering from CRSwNP are often as affected by chronic asthma [17,18]. Given that CRS affects 3-15% of the population in the US and Europe, with a prevalence of 2.7% in Sweden [19–21], the major focus of this study was therefore to determine whether ACE2 expression is altered in patients with CRSwNP. To achieve this, RT-qPCR was used to evaluate the expression of the *ACE2* mRNA transcript in the nasal epithelium of patients with CRSwNP. To determine whether there were any differences in ACE2 expression, tissue samples from patients were collected from non-polyp ‘healthy’ tissue and adjacent polyp tissue, as well as from control individuals without CRSwNP.

Recent studies have demonstrated that ACE2 is highly expressed in the nasal epithelial cells, highlighting the importance of the upper airway tissues as the primary target site of SARS-CoV-2 [9–11]. To determine *ACE2* mRNA expression in the nasal epithelium of CRSwNP patients, we performed RT-qPCR on total RNA extracted from concha inferior or middle and polyp mucosa in a total of 16 patients and 18 non-CRSwNP controls. We found that *ACE2* mRNA expression was significantly reduced in the nasal mucosa of non-polyp ‘healthy tissue’ of CRSwNP patients compared to that of non-CRSwNP controls, and in turn *ACE2* expression in non-polyp ‘healthy tissue’ was significantly reduced compared to that of their adjacent polyp tissue (Figure 1). However, no significant differences in expression of the *ACE2* gene transcript were observed between healthy CRSwNP controls and polyp tissues of CRSwNP patients (Figure 1). Taken together, these observations indicate that patients with CRSwNP may have a lower risk of SARS-CoV-2 infection due to lower expression levels of *ACE2* gene.

**Figure 1.**
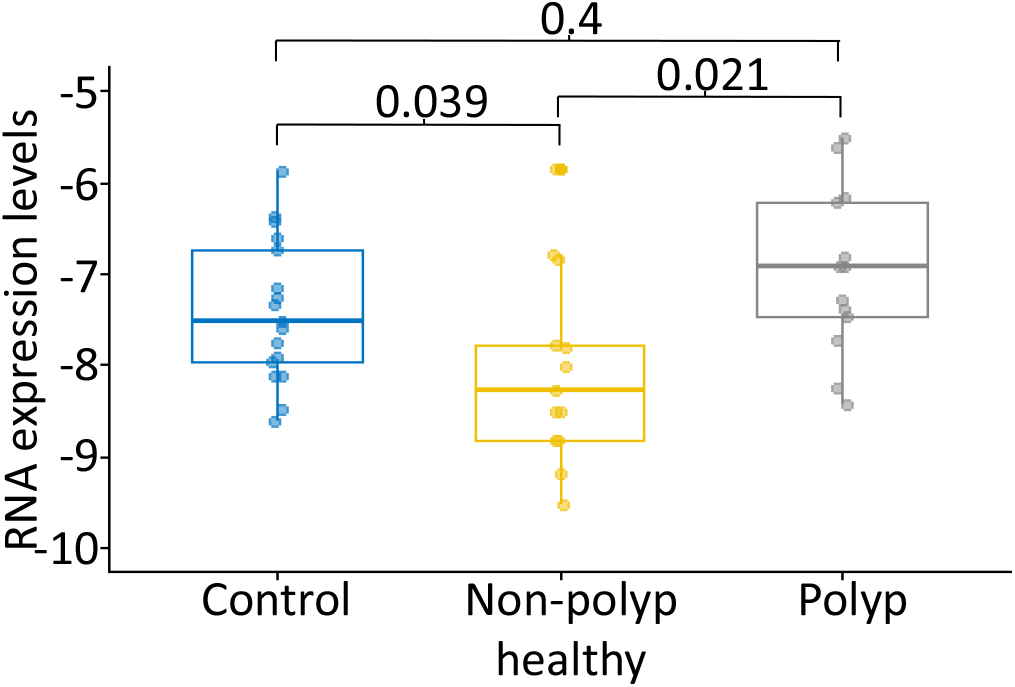
Nasal gene expression of *ACE2* is altered in patients with chronic rhinosinusitis with nasal polyps. Gene expression of *ACE2* in the inferior or middle nasal concha of non-CRS controls (n=18), non-polyp healthy tissue and adjacent polyp tissue of CRswNP patients (n=16). The data presented shows lower *ACE2* gene expression in the nasal mucosa of non-polyp healthy tissue compared to that of adjacent polyp tissue and non-CRS controls.

Whilst an age-dependant gene expression of ACE2 has recently been reported [16], the impact of gender on ACE2 gene expression remains poorly understood. Here we found that *ACE2* mRNA levels significantly differ between female CRSwNP patients compared to that of males. More specifically, we found that *ACE2* mRNA expression was significantly reduced in the nasal mucosa of female non-polyp ‘healthy tissue’ of CRSwNP patients compared to that of non-CRSwNP female controls, whereas no significant differences were observed in males (Figure 2 and Table 1). Moreover, *ACE2* mRNA expression was significantly decreased in non-polyp ‘healthy tissue’ of female CRSwNPs patients compared to their adjacent polyp tissue, where no significant differences were observed in males (Figure 2 and Table 1). Taken together, our results indicate a gender bias of ACE2 expression in CRSwNP patients, where females with CRSwNPs have a lower expression of nasal *ACE2* gene expression.

**Table 1.** **Statistical comparison of nasal *ACE2* gene expression in males and females of patients with chronic rhinosinusitis with nasal polyps**

| Sex | T-test | P-value summary |
| --- | --- | --- |
| Female | Control vs non-polyp healthy | 0.00093 |
|  | Control vs polyp | 0.0057 |
|  | Non-polyp healthy vs polyp | 0.93 |
| Male | Control vs non-polyp healthy | 1 |
|  | Control vs polyp | 0.51 |
|  | Non-polyp healthy vs polyp | 0.47 |

**Figure 2.**
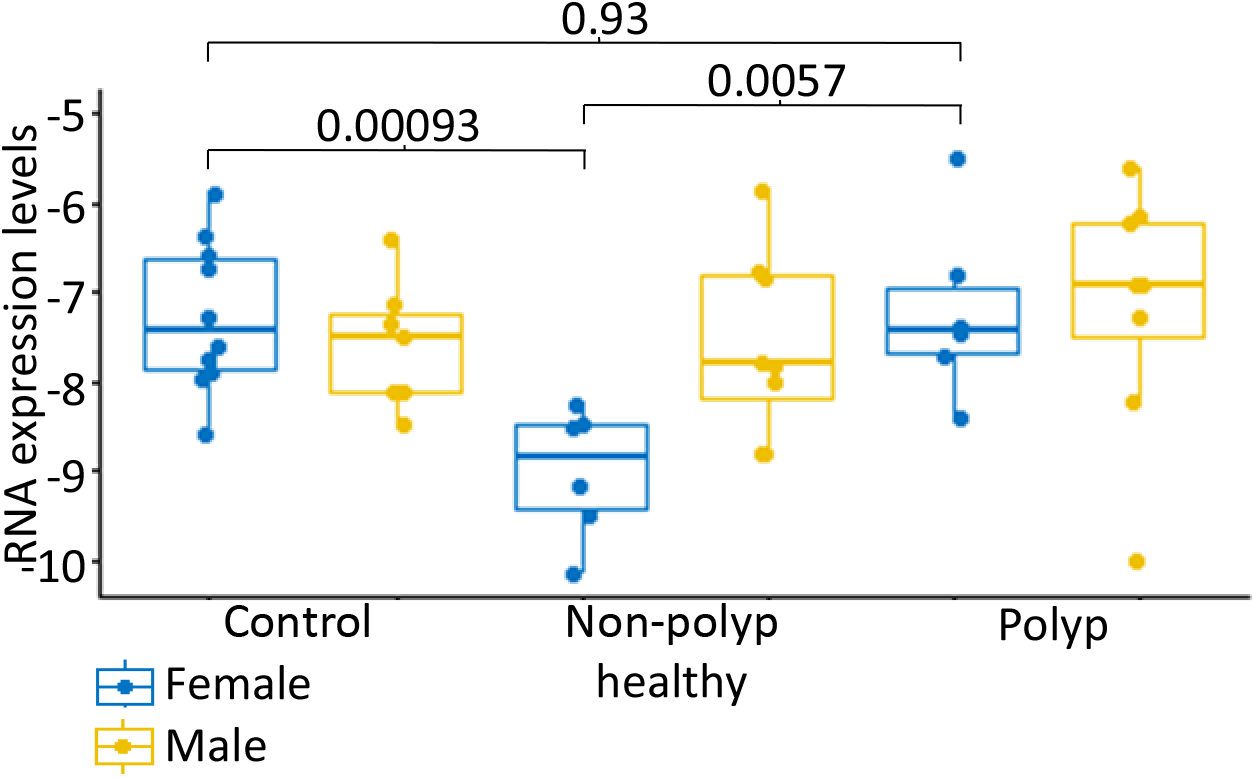
Sex-dependent differences of nasal *ACE2* gene expression in patients with chronic rhinosinusitis with nasal polyps. Gene expression of *ACE2* in the inferior or middle nasal concha of non-CRS controls (n=18), non-polyp healthy tissue and adjacent polyp tissue from CRswNP patients (n=16). The data presented shows lower *ACE2* gene expression in the nasal mucosa of females in both non-polyp healthy tissue and adjacent polyp tissue compared to non-CRS controls. No significant differences in *ACE2* gene were observed in males.

The mechanisms driving altered susceptibility to COVID-19 remains a crucial question for the field. The high expression of ACE2 has been suggested to be a potential risk factor for virus invasion, therefore understanding the association between CRS and ACE2 expression is particularly important for the management of upper airway conditions which may help to predict the patient’s susceptibility to COVID-19. In this study we have demonstrated that ACE2 is differentially expressed in the nasal tissue of patients with CRSwNPs compared to healthy individuals, where *ACE2* gene expression is significantly reduced in non-polyp ‘healthy tissue’ of CRSwNP patients but not adjacent polyp tissue. These changes seem to occur in a sex dependent manner where changes to the *ACE2* gene transcript is only observed within female patients in a more profound manner. This is the first study to report a lower *ACE2* gene expression in the nasal tissue of females CRSwNPs patients compared to that of CRSwNPs males. Taken together, our results thereby suggest that CRSwNP patients, especially females that have a reduction in nasal *ACE2* gene expression, may be associated with a lower risk to be infected with SARS-CoV-2.

Whilst ACE2 gene expression has previously been examined in CRS patients, the current study confirms previous observations and reports a significantly lower *ACE2* gene expression in the nasal mucosa of patients with CRSwNPs compared to healthy individuals [22–24]. However, this is the first study to report a lower *ACE2* gene expression in CRSwNP patients in a Swedish cohort. Interestingly, similar ACE2 gene expression profiles have been reported in other airway conditions, including asthma and chronic rhinitis, which have both been associated with significant reductions in ACE2 gene expression in airway epithelial cells [25–27]. CRS is often associated with asthma [17,18,28,29], and diseases of the upper airways with a decrease in ACE2 expression may thereby play an important role in lowering the risk of infection with SARS-CoV-2. In keeping with this notion, a recent report has shown that CRS prevalence in hospitalised COVID-19 patients in Wuhan, China, was 6.1%, which is somewhat lower than the prevalence of the disease in the general population of China (8%), and that CRS comorbidity is not associated with the severity of COVID-19 [30]. Further, Chhiba et al., 2020 have reported that COVID-19 patients with asthma and rhinosinusitis present a lower risk of hospitalisation than patients without these conditions [31]. However, whilst a direct link between decreased ACE2 expression and decreased susceptibility to acquisition of SARS-CoV-2 has been reported, there is also evidence to suggest that a downregulation of ACE2 could potentially worsen COVID-19 symptoms. Lung involvement is the most severe complication of COVID-19, often evolving into acute respiratory distress syndrome [3]. Previous reports have suggested that ACE2 plays a protective role in acute respiratory distress syndrome, where studies have shown that loss of ACE2 expression exacerbates symptoms in different models of acute lung injury [32,33]. Lastly, although a reduction in ACE2 gene expression has been reported in CRS, conflicting observations exist regarding ACE2 protein expression in CRS patients. For example, whilst using immunohistochemistry to evaluate ACE2 protein expression within the nasal cavity of CRS patients, no differences were observed in ACE2 protein expression compared to that of healthy controls [34,35]. Contradictory to this, Wang et al., 2020 reported a reduction in ACE2 protein expression in CRS patients both with and without nasal polyps [22]. Therefore, more studies are warranted to determine exactly whether the expression of ACE2 in the upper airway patients could influence the risk of infection with SARS-CoV-2.

A striking feature of the COVID-19 outbreak is the difference in morbidity and mortality between the sexes. The infection rate between males and females seem to vary depending on geographical location, albeit the Global Health 50/50 project has found similar numbers of COVID-19 cases in men and women in 18 different countries (https://globalhealth5050.org/the-sex-gender-and-covid-19-project/). Whilst the infection rate between men and females are similar, a much higher mortality and more severe presentation of the disease is observed in men. For example, in Chinese confirmed cases the death rate among men is 4.7% compared with 2.8% for women, where in Italy the death rate among men is 16.6% compared with 9.1% in women [36,37]. More recently, a meta-analysis study of 39 reports, including 206,128 reported cases, confirmed a sex bias where male patients with COVID-19 have more than double the rate of requiring intensive treatment and have a significantly higher mortality rate than females [38]. Lastly, in Sweden, the percentage of confirmed COVID-19 cases is slightly lower in males (46.32%) compared to that of females (53.68%) the mortality rate in males is 3.39% compared to 2.44% in females (https://globalhealth5050.org/the-sex-gender-and-covid-19-project/). In the current study, we report a significant decrease in the nasal *ACE2* gene expression in only female CRSwNP patients. A lower ACE2 gene expression in females could potentially explain part of the gender disparity observed in COVID-19, it however remains unclear whether there are any general gender differences in ACE2 gene expression. Recently, Cai 2020 analysed four large-scale bulk transcriptomic datasets of normal lung tissue and two single-cell transcriptomic datasets and did not find any sex differences in ACE2 mRNA expression between males and females [39]. In contrast to this, Shastri et al., 2020 observed that ACE2 was more highly expressed in testicular cells compared to ovarian tissue, raising the possibility that testicular ACE2 reservoirs may play a role in viral persistence in males. Lastly, a higher activity, but not expression, of ACE2 has been reported in male mouse kidneys compared to females [40]. To our knowledge, our study herein is the first report on a gender difference in the nasal ACE2 gene expression.

The recruitment and activation of eosinophils, mast cells and innate lymphoid cells have been shown to contribute to the chronic inflammatory state in CRS [41,42]. Significantly higher levels of the prominent Th2 cytokines IL-4, IL-5 and IL-13 have been reported in nasal polyp tissue from CRSwNP patients which coincided with reduced ACE2 expression [24]. Interestingly, IL-4 has been shown to modulate ACE2 gene expression in SARS-CoV-2 infected cells, where treatment of Vero E6 cells with IL-4 decreased the susceptibility of these cells to be infected with SARS-CoV-2 and decreased the expression of ACE2 [43]. In a similar fashion, IL-13 has additionally been shown to modulate ACE2 gene expression in airway epithelial cells in asthma [44]. However, it is important to take into account that all CRSwNP patients that participated in our study were treated with corticosteroids locally in their nasal cavity, which might have served as a confounding factor in our study. A recent report suggested that in asthmatic subjects, ACE2 expression is significantly lower in patients who are on inhaled corticosteroids compared to those who are not [45]. Similarly, corticosteroid administration was shown to reduce the expression of ACE2 in airway epithelial cell cultures from COPD patients [46]. Given that there has additionally been evidence to suggest that ACE2 expression may directly be regulated by exposure to corticosteroids [24], it is therefore likely that the local corticosteroid administration impacts ACE2 gene expression in CRSwNP patients. The beneficial effect of short-term treatment with corticosteroids in severe covid-19 patients could, at least in part, be explained by the role of corticosteroid in lowering ACE2 expression. Nevertheless, the underlying mechanism of the observed beneficial effect of treatment with corticosteroids in CRS and other airway disorders in suppression of ACE2 expression, and reduction of SARS-CoV-2 infection remains to be established.

In summary, this study reports that the gene expression of the SARS-CoV-2 entry factor *ACE2* is differentially expressed in the nasal mucosa of patients with CRSwNPs in a Swedish cohort. We also report a sex-dependant difference in nasal *ACE2* expression, where significantly lower levels of the *ACE2* transcript were detected in the nasal mucosa of only female CRSwNP patients. Further studies are warranted to elucidate the underlying mechanism of the altered ACE2 expression in CRSwNPs patients nasal mucosa, and its potential impact on the acquisition of SARS-CoV-2 infection and the disease severity of COVID-19.

## Methods

### Patients and sample collection

A total of 35 individuals were included in this study, divided into two groups: CRSwNP patients (n=16) and healthy controls (n=18). Control samples were obtained from individuals aged 40 to 84, with a median age of 60 years and without any history of sinus disease. Patients with CRSwNP ranged in age from 35 to 73, with a median age of 60 years. The diagnosis of CRS was based on the definition of the European Position Paper on Rhinosinusitis and Nasal Polyps 2012 guidelines [47]. Cutting through biopsies were taken from concha inferior or middle from CRSwNP patients and controls under narcosis with Rudolf Weil-Blakesley 3mm forceps. An additional biopsy was taken from the polyp tissue of CRSwNP patients. All biopsies were immediately put in RNA later preservative fluid (ThermoFisher, San Diego, CA, USA). All patients were on oral corticosteroids for the treatment of CRS as recommended by the current Rhinosinusitis and Nasal Polyps 2012 guidelines. The study was carried out in accordance with the Declaration of Helsinki and was approved by the Local Ethics Committee in Gothenburg, Sweden (Date for approval 2012-12-30, Dnr 829-12).

### RT-qPCR analysis

RNA Total RNAs from biopsies were isolated using the Kingfisher total RNA extraction kit (ThermoFisher, San Diego, CA, USA) according to the manufacturer’s protocol. cDNAs were synthesized from total RNAs by using a cDNA synthesis kit (Vilo cDNA kit/ ThermoFisher, San Diego, CA, USA), according to the manufacture’s protocol. RT-qPCR was performed on the quantstudio12Kflex instrument (ThermoFisher, Life Technologies, CA, San Diego, USA) using TaqMan Gene Expression Master Mix (ThermoFisher, San Diego, CA, USA) with technical duplicates. Primer and probe sets for ACE2 (Hs01085333_m1 and Hs00222343_m1) were purchased from ThermoFisher, San Diego, CA, USA. Relative gene expression was calculated using the comparative crossing threshold (*C*_*T*_) method, where the geometrical means of YWHAZ (Hs01122445_g1), HPRT1 (Hs02800695_m1) and GUSB (Hs00939627_m1) were used as a reference for housekeeping genes.

### Statistical analysis

Statistical analysis for ACE2 expression was performed using R Version 3.6.2, and data are presented as medians and interquartile range. A general linear model using age or sex as covariates was used to compare ACE2 expression of controls vs non-polyp ‘health tissue. Wilcoxon matched-pairs signed rank test was used for the analysis of paired data (i.e. non-polyp ‘healthy’ tissue vs adjacent polyp tissue). Differences were considered statistically significant at *P*-value < 0.05.

## Data Availability

The data that support the findings of this study are available from the corresponding author (ATN) upon reasonable request.

## Acknowledgements

The authors are grateful to all patients and healthy individuals who participated in this study. This study has been supported by grants from the Hospital of Skaraborg, Öronkliniken vid KSS research fund, the Health and Medical Care Committee of the Regional Executive board, Region Västra Götaland, the Foundation of Acta Oto-Laryngologica, the ENT Foundation and VBG Group Centre for Asthma and Allergy Research.

## Notes

### Competing Interest Statement

The authors have declared no competing interest.

### Funding Statement

This study has been supported by grants from the Hospital of Skaraborg, Oronkliniken vid KSS research fund, the Health and Medical Care Committee of the Regional Executive board, Region Vastra Gotaland, the Foundation of Acta Oto-Laryngologica, the ENT Foundation and VBG Group Centre for Asthma and Allergy Research.

### Author Declarations

The study was carried out in accordance with the Declaration of Helsinki and was approved by the Local Ethics Committee in Gothenburg, Sweden (Date for approval 2012-12-30, Dnr 829-12).

